# Genetic Diversity of the *Plasmodium falciparum* Reticulocyte Binding protein Homologue-5 which is a potential Malaria Vaccine Candidate: Baseline data from areas of varying malaria endemicity in Mainland Tanzania

**DOI:** 10.1101/2024.09.20.24314052

**Authors:** Angelina J. Kisambale, Beatus M. Lyimo, Dativa Pereus, Salehe S. Mandai, Catherine Bakari, Gervas A. Chacha, Ruth B. Mbwambo, Ramadhan Moshi, Daniel A. Petro, Daniel P. Challe, Misago D. Seth, Rashid A. Madebe, Rule Budodo, Sijenunu Aaron, Daniel Mbwambo, Abdallah Lusasi, Stella Kajange, Samwel Lazaro, Ntuli Kapologwe, Celine I. Mandara, Deus S. Ishengoma

**Affiliations:** National Institute for Medical Research, Dar es Salaam, Tanzania; Nelson Mandela African Institution of Science and Technology, Arusha, Tanzania; Muhimbili University of Health and Allied Sciences, Dar es Salaam, Tanzania; University of Dar es Salaam, Dar es Salaam, Tanzania; National Institute for Medical Research, Tanga Research Centre, Tanga, Tanzania; National Malaria Control Programme, Dodoma, Tanzania; President’s Office, Regional Administration and Local Government, Dodoma, Tanzania; Directorate of Preventive Services, Ministry of Health, Dodoma, Tanzania; Department of Biochemistry, Kampala International University in Tanzania, Dar es Salaam, Tanzania

**Keywords:** Malaria, malaria Vaccine, Pfrh5, Plasmodium falciparum, Genetic diversity, Mainland Tanzania

## Abstract

**Background:** The limited efficacy of the two malaria vaccines, RTS,S/AS01 and R21/Matrix M, which were recently approved vaccines by the World Health Organization, highlights the need for alternative vaccine candidate genes beyond these pre-erythrocytic-based vaccines. *Plasmodium falciparum* Reticulocyte Binding Protein Homologue 5 (*Pfrh5)* is a potential malaria vaccine candidate, given its limited polymorphism compared to other parasite’s blood stage antigens. This study evaluated the genetic diversity of the *Pfrh5* gene among parasites from regions with varying malaria transmission intensities in Mainland Tanzania, to generate baseline data for this potential malaria vaccine candidate.

**Methods:** This study utilized secondary data of 697 whole-genome sequences from Mainland Tanzania, which were generated by the MalariaGEN Community Network. The samples which were sequenced to generate the data were collected between 2010 and 2015 from five districts within five regions of Mainland Tanzania, with varying endemicities (Morogoro urban district in Morogoro region, Muheza district in Tanga region, Kigoma-Ujiji district in Kigoma region, Muleba district in Kagera region, and Nachingwea district in Lindi region). The genetic diversity of the *Pfrh5* gene was assessed using different genetic metrics, including Wright’s fixation index (F_ST_), Wright’s inbreeding coefficient (Fws), Principal Component analysis (PCA), nucleotide diversity (***π***), haplotype network, haplotype diversity (Hd), Tajima’s D, and Linkage disequilibrium (LD).

**Results:** Of the sequences used in this study (n=697), 84.5% (n = 589/697) passed quality control and 313 (53.1%) were monoclonal, and these monoclonal sequences were used for haplotype diversity and haplotype network analysis. High within-host diversity (Fws <0.95) was reported in Kigoma-Ujiji (60.7%), Morogoro urban (53.1%), and Nachingwea (50.8%), while Muleba (53.9%) and Muheza (61.6%) had low within host diversity (Fws≥0.95). PCA did not show any population structure across the five districts and the mean F_ST_ value among the study populations was 0.015. Low nucleotide diversity values were observed across the study sites with the mean nucleotide diversity of 0.00056. A total of 27 haplotypes were observed among the 313 monoclonal samples. The *Pf*3D7 was detected as Hap_1, and it was detected in 16/313 (5.1%) sequences, and these sample sequences were from Muheza (62.5%, n=10/16), Kigoma-Ujiji (18.8%, n=3/16), and Muleba (18.8%, n=3/16). Negative Tajima’s D values were observed among the parasite populations in all the study sites.

**Conclusion:** In this study, we observed low levels of polymorphism in the *pfrh5* gene, as it exhibited low nucleotide and haplotype diversity, a lack of population structure and negative Tajima’s D values as signatures of purifying selection. This study provides an essential framework of the diversity of the *Pfrh5* gene to be considered in development of the next generation malaria vaccines. Robust and intensive studies of this and other candidate genes are required for characterization of the parasites from areas with varying endemicity, and are crucial to support the prioritization of the *Pfrh5* gene for potential inclusion in a broadly cross-protective malaria vaccine.

## Background

Despite substantial efforts in investing in interventions to control and eliminate malaria, the disease continues to pose a significant global health challenge [1]. The World Health Organization (WHO) reported about 249 million malaria cases and 608,000 deaths in 85 malaria-endemic countries in 2022, with most of the cases (93.6%) and deaths (95.4%) from the WHO African region [2] ; and over 96.0% of the cases were due to *Plasmodium falciparum*. According to the 2023 World Malaria Report, Tanzania was among the 11 countries with the highest number of malaria deaths and cases in Sub Saharan Africa (SSA); it accounted for 3.2% of the global cases and 4.4% of global malaria deaths [2]. Over 93.0% of the population in Tanzania lives in areas where transmission occurs, and the entire population of Mainland Tanzania is considered at risk of malaria. However, the transmission rates and burden vary among and within regions, with high transmission in western, northwestern and southern regions [3].

Despite the efforts deployed in the past 2.5 decades, malaria control and elimination still faces various challenges including the key biological threats; insecticide resistance by mosquitoes [4], resistance of parasites to antimalarials [5,6], emergence of parasites with deletions of histidine rich protein 2/3 genes [7](these cannot be detected by rapid diagnostic tests), and invasive vector species; *Anopheles stephensi* [8,9]. Also there are non-biological factors such as climate change [10] and health system as well as reduced funding [11] which significantly reduce the impact of the interventions which have been deployed to control and eventually eliminate malaria. The control of malaria also faces challenges which are attributed to the complex life cycle of *Plasmodium* parasites, as the parasite undergoes several stages of development in both human and vector hosts [12–14]. To tackle the parasite-related challenges, it is crucial to incorporate supplementary measures, such as vaccines, into the current approaches for controlling and eliminating malaria. The effectiveness of vaccines in the global effort to control, eliminate and eradicate other serious diseases like polio, COVID-19, and smallpox underscores their potential to enhance malaria control, elimination and eventual eradication [15,16].

Recently, WHO approved two malaria vaccines, RTS,S/AS-01 in 2021 and R21/Matrix-M in 2023 for malaria prevention in children from countries/areas with moderate to high malaria burden [17]. Both vaccines are subunit vaccines, targeting the *P. falciparum* circumsporozoite protein (*Pf*CSP) [18]. This protein is expressed in pre-erythrocytic stages on the surface of sporozoites and is responsible for hepatocyte invasion [17]. Studies have demonstrated that the vaccines exhibit greater efficacy against parasites with alleles matching the vaccine strains compared to diverse alleles in the targeted populations [18]. Comparative diversity studies of the 3D7 *Pfcsp* gene revealed that only 0.2% to 5.0% of the vaccine strains align with the global *Pfcsp* gene [16]. Studies from different malaria-endemic countries including Tanzania [19], Ghana[18] China [20] and Myanmar [21], have demonstrated a significant level of genetic diversity in the *Pfcsp* gene, hence standing as one of the challenges for the efficacy of these vaccines [7]. Polymorphisms in genes related to malaria vaccine candidates are one of the key factors hindering the development of other malaria vaccines and needs to be considered when selecting and optimizing any potential malaria vaccine candidate [22,23]. Therefore, there is an urgent need to utilize the recently generated global genomic data of malaria parasites to identify highly conserved vaccine candidates, which will facilitate the development of highly efficacious vaccines that will be critical in the ongoing malaria elimination and eradication efforts.

The *P. falciparum* reticulocyte binding protein homologue 5 (*Pfrh5*) complex stands out as a crucial target for developing an effective vaccine targeting the blood stage[24]. *Pfrh5*, a 63 kDa protein is encoded by the gene PF3D7_0424100, and plays a key role in invasion of erythrocytes by the merozoites. The *pfrh5* forms a pentameric complex containing *P. falciparum* thrombospondin-related apical merozoite protein, *P. falciparum* cysteine-rich protective antigen and PfRH5-interacting protein post-release from the rhoptries. This process facilitates *Pfrh5* expression on the merozoite surface during invasion, where its interaction with the receptor basigin (BSG) also known as CD147 via hydrogen bonds is vital for erythrocyte invasion [25,26]. Antibodies targeting the PfRH5-BSG invasion pathway have demonstrated significant inhibition of invasion of erythrocytes [27]. Studies have also highlighted the effectiveness of *Pfrh5* antibodies in inhibiting the growth of various *P. falciparum* strains, surpassing other antigens on vaccine development platforms [24,28,29]. Moreover, *Pfrh5* exhibits relatively low genetic variation, with a limited number of non-synonymous mutations which have been identified in this gene [30,31]. This conservation with minimal polymorphism, sets *Pfrh5* apart from other prominent subunit vaccine candidates like apical membrane antigen-1 [32] and merozoite surface protein-1 (MSP1)[33]. Before proceeding with vaccine development based on *Pfrh5*, a thorough investigation and characterization of *Pfrh5* genetic diversity in regions with varying malaria endemicity and heterogeneous transmission patterns, such as Tanzania, are imperative [34]. Therefore, this study was conducted to evaluate the genetic diversity within the *Pfrh5* gene isolated from parasites collected in areas exhibiting varying levels of transmission intensities in Mainland Tanzania, to generate baseline data of this potential malaria vaccine candidate.

## Methods

### Study design and sampling

The study utilized data which were retrieved from an open dataset of the *P. falciparum* genome variation of the Genomic epidemiology of malaria network (MalariaGEN Pf7) [35]. The collection of samples was conducted as part of the Pathogen Diversity Network Africa baseline surveys across 15 countries in SSA [36]. The samples collection was done from 2010 to 2015 through different studies including, the therapeutic efficacy studies (TES) of children aged 6 months to 10 years, cross section survey (CSS) of patients aged 6 months and above, and longitudinal and cross-sectional birth cohort studies. The TES were conducted at Mkuzi health centre in Muheza district (Tanga region), Muheza designated district hospital (DDH) also in Muheza district (Tanga region), and Ujiji health centre in Kigoma-Ujiji district (Kigoma region) as previously described [37–39]. The CSS were conducted at Rubya DDH in Muleba district (Kagera region), Mkuzi health centre in Muheza district (Tanga region) and Nachingwea district hospital in Nachingwea district (Lindi region) [40]. The longitudinal and cross-sectional birth cohort studies were conducted at Muheza DDH (Tanga region) and Morogoro regional hospital in Morogoro urban district (Morogoro region) through the Mother and offspring malaria study as described earlier [41,42]. Based on the 2020 stratification of malaria burden; Muheza and Morogoro urban were classified as areas with moderate transmission of malaria while Nachingwea, Muleba and Kigoma-Ujiji were areas with high transmission intensity [43]. Similar data have also been used for studies of CSP polymorphism and evidence of selection in Tanzania [19].

The analysis involved a total of 697 samples that were collected as whole blood from the five districts in the five regions in Mainland Tanzania. The number of samples from each site included 324 samples in Muheza district, 34 in Morogoro urban, 199 in Kigoma-Ujiji, 79 in Nachingwea and 61 samples in Muleba district (**Table 1**). Parasite genomic DNA was extracted using QIAamp DNA blood mini kits (Qiagen GmbH, Hilden, Germany) and genomic data were generated by whole genome sequencing (WGS) using illumina short reads which were done through the *P. falciparum* Community Project of MalariaGEN at the Wellcome Sanger Institute, UK as described earlier [35].

**Table 1:** Details of samples used for this study.

| Study site | Retrieved sequences | Passed filtering score | Monoclonal(%) | Polyclonal(%) |
| --- | --- | --- | --- | --- |
| Morogoro urban* | 34 | 32 | 15(46.9) | 17(53.1) |
| Muheza** | 324 | 297 | 182(61.3) | 115(38.7) |
| Muleba** | 61 | 52 | 27(51.9) | 25(48.1) |
| Nachingwea** | 79 | 65 | 32(49.2) | 33(50.8) |
| Kigoma-Ujiji*** | 199 | 143 | 57(39.9) | 86(60.1) |
| **Total** | **697** | **589** | **313(53.1)** | **276(46.9)** |

\*Sample collection in Morogoro urban was done through the Mother and offspring Malaria study which was also conducted at Muheza DDH [41,42].
\*\*Samples from Muheza were collected through two therapeutic efficacy studies and a cross-sectional study which also covered Nachingwea and Muleba [37,38,40].
\*\*\*Sample collection in Kigoma-Ujiji was done in a therapeutic efficacy study as previously reported [38,39].

### Data retrieval and sequence acquisition

WGS of 697 *P. falciparum* and sample information from each study site were mined from the database of the MalariaGEN *Plasmodium falciparum* Community (Pf7k) project in variant call format (VCF). The *Pfrh5* gene information including genomic location and genomic length were obtained from the PlasmoDB database http://www.plasmodb.org/. The gene was extracted from chromosome four at position 1082005-1084464 using bcftools [44,45].

Before progressing with downstream data analysis, the sequences were filtered to include only biallelic data with a variant quality score log odds (VQSLOD) of ≥1. The biallelic SNPs that passed quality filter scores were used for further analysis [46]. To obtain the FASTA format sequences from the processed VCF files, the Pf3D7 reference genome (Pf3D7_04_03 1 1200490) was indexed using Samtools v 1.18, and then Picard v 3.1.0 tool was used to create a sequence dictionary for the reference genome (https://broadinstitute.github.io/picard/). Subsequently, the Genome analysis toolkit (GATK v 4.4.0.0) was used to index the processed VCF files and generate FASTA files containing the alternative reference for the processed VCF files [47]. The FASTA files were then merged into one major file for subsequent downstream analysis. Further analysis of genetic metrics was done on a DnaSP v 6.12 software and R workflow v.4.4.0. The data analysis processes are summarised in **Fig.2**.

**Figure 1:**
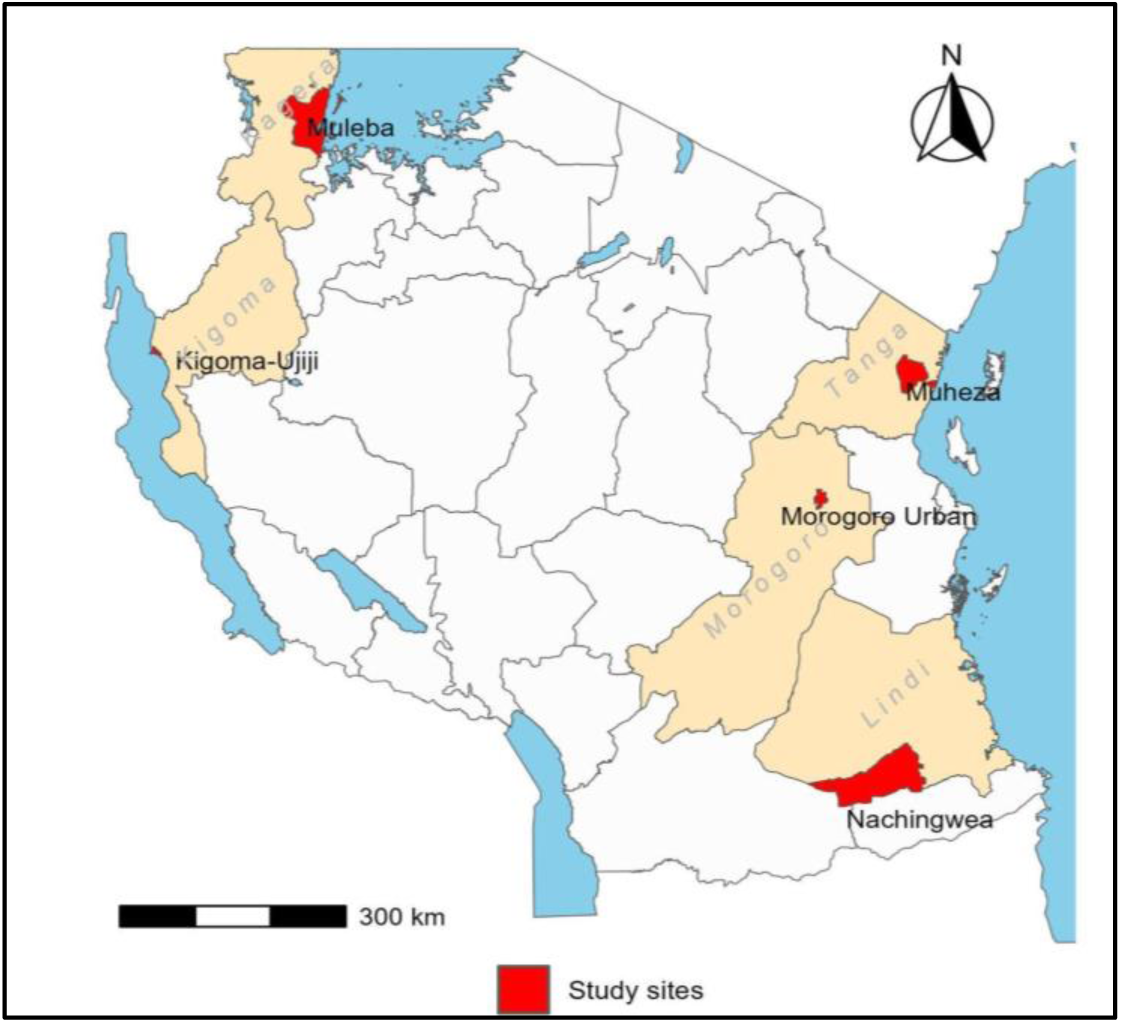
Map of Tanzania showing the five sites where sampling was undertaken (red colour).

**Figure 2:**
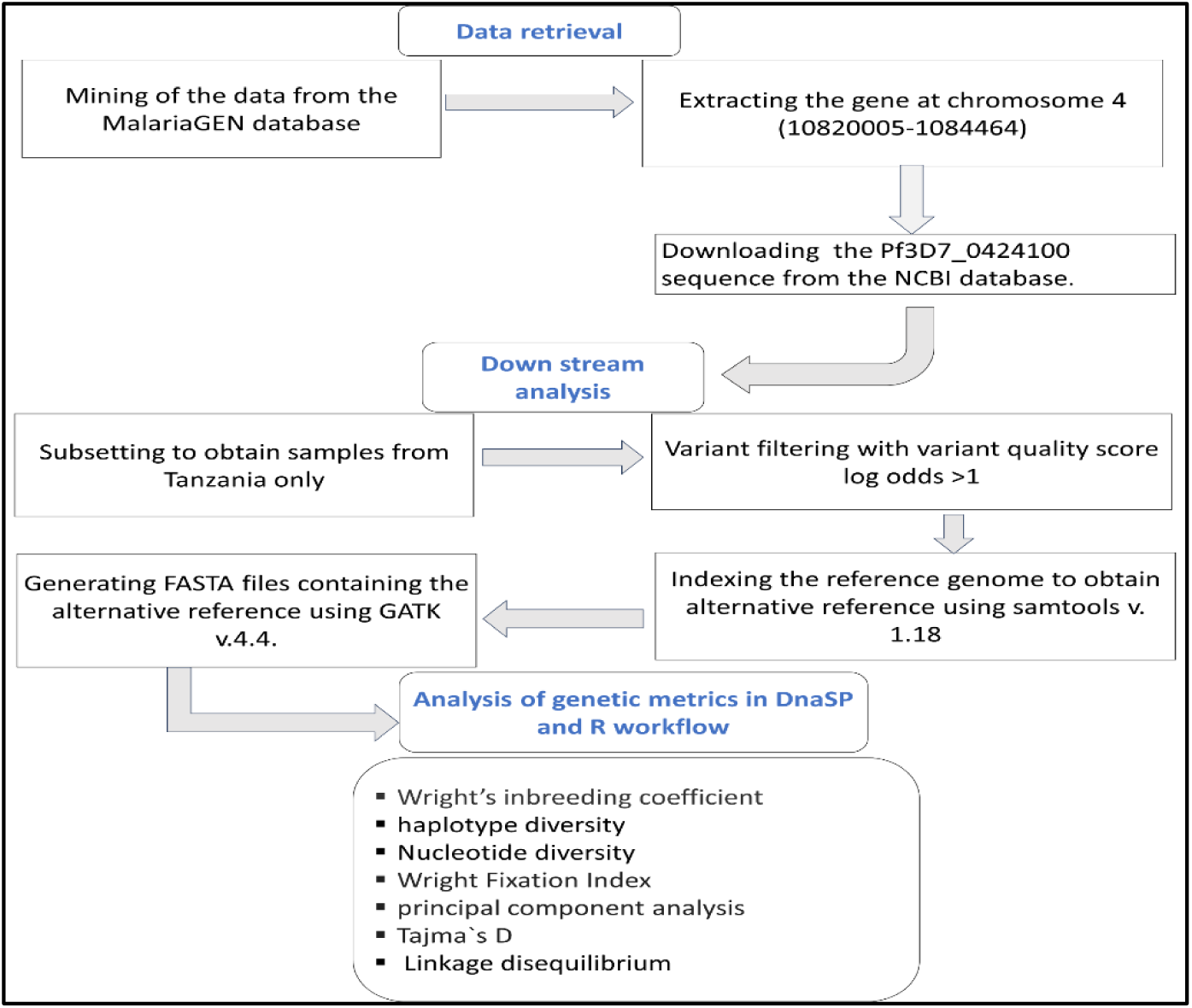
Summary of the steps involved in data retrieving and downstream analysis of the Pfrh5 DNA sequences.

#### Population genetics analysis

##### Within host-parasite diversity

Within-host diversity was evaluated by estimating Wright’s inbreeding coefficient (Fws) for each population using the “moimix” package in R v 4.4.0[48]. The Fws metric measured the degree of polyclonal infections, assessing the within-host diversity of *Plasmodium* in relation to local population diversity [28]. Fws ranges from 0 to 1, a low Fws value indicates high degree of genetic diversity within the parasite population compared to the overall population. Samples with a high Fws value (≥0.95) were classified as monoclonal (single strain) infections, while those with a low Fws value (<0.95) were regarded as polyclonal infections (mixed strain infections) [49,50]. A Pearson chi-square test was conducted in R v.4.4.0 to assess and determine the differences in Fws estimates among the study populations, with a standard threshold of p <0.05 indicating statistical significance.

##### Genetic diversity within parasite populations

FASTA DNA sequences were generated from the VCF files using the G.A.T.K v4.4.0.0. Different genetic metrics were used to assess the diversity of the *Pfrh5* gene within each parasite population using DnaSP v.6.12 software. These genetic metrics included the number of segregating sites (S), referred to as the position within the gene sequence where there are variations between alleles observed in a population, the number of haplotypes (H: A set of DNA variants along a chromosome that tend to be inherited together), and haplotype diversity (Hd); a measure of the variety of haplotypes present within a population. Others included, singleton variable sites (Sn); for mutations appearing only once among the sequences, and Parsimony informative sites; (sites containing at least two types of nucleotides and at least two of them occurring with a minimum frequency which is required for evolutionary changes of a genome). Nucleotide diversity was used to measure the average number of nucleotide differences per site between any two randomly chosen DNA sequences within a population estimated as (***π***). Violin plots were used to visually assess the median differences of nucleotide diversity between the populations, implemented in ggplot2 packages in R workflow. To examine the genetic connectivity among *Pfrh5* haplotypes across the five districts, the haplotype networking of 313 *Pfrh5* monoclonal sequences (53.1%) was analysed using NETWORK version 10.2, employing the Median-joining algorithm [51]. The sequence of PF3D7_0424100, a reticulocyte binding protein homologue 5 (*Pf*3D7), was downloaded from the NCBI database https://www.ncbi.nlm.nih.gov/gene/812437 and used to compare its variation and clustering with those of natural parasite populations.

##### Population structure and differentiation

To assess gene flow between parasite populations, genetic differentiation was estimated using the Wright Fixation Index (F_ST_) [43] using the PopGenome package in R v 4.4.0 workflow. The F_ST_ measures population differentiation due to genetic structure, and values of <0.05 indicate minimal population differentiation or gene flow between pairs of populations [52]. Violin plots were also generated to visualize the differences on F_ST_ values among the study populations using the ggplot2 packages in R. Principal component analysis(PCA); a linear technique used for data visualization [53] was also conducted to assess the population structure and it was implemented in R version 4.4.0.

##### Evidence of selection and genetic recombination

The neutrality tests were performed to determine whether the *Pfrh5* gene is under balancing or purifying selection. This was done using Tajmàs D statistical test to test the departure of a gene from neutrality theory based on allele frequency distribution in the gene. The analysis was performed in sliding windows with a window length of 100 bp and a step size of 25, utilizing the total number of mutations and excluding sites with gaps in a DnaSP v. 6.12 Software [54,55]. To validate the observed values of signature of selection, Fu and Li’s D and F test statistics were also assessed. Fu and Li’s D test statistic calculated the differences between the observed number of singletons (mutations appearing only once among the sequences), and the total number of mutations while the Fu and Li’s F test statistic considered the differences between the number of singletons and the average number of nucleotide differences between pairs of sequences[56,57]. Additionally, Linkage disequilibrium (LD) was computed considering all the polymorphic sites to assess the level of non-random association between alleles. Genetic association between polymorphic sites and the effect of intragenic recombination on sequence polymorphism were assessed on the polymorphic sites using (Zns) [58] and the ZZ [59] statistics, respectively.

## Results

### Baseline information

A total of 697 WGS were retrieved from the MalariaGEN Pf7 database. Of the retrieved sequences, 84.5%(n = 589/697) passed quality filtering scores and were subsequently used for downstream analysis. Among the sequences which passed the filtering scores, 50.4% (n = 297/589) were from Muheza district while Kigoma - Ujiji had 24.3% (n = 143/589), and the rest were from the three remaining sites, with 11.0% (n = 65/589) from Nachingwea, 8.8% (n = 52/589) from Muleba and 5.4% (n = 32/589) from Morogoro urban district. Of the successfully retrieved sequences 53.1% (n= 313/589) had monoclonal infections and were used for haplotype diversity and haplotype network analysis.

### Within-host genetic diversity

The degree of genetic diversity within each population was estimated through the wright’s inbreeding coefficient (Fws), and showed that the majority of the samples from Kigoma -Ujiji (60.1%, n = 86/143), Morogoro urban (53.1%%, n = 17/32) and Nachingwea (50.8%, n = 33/65) had polyclonal infections (Fws<0.95). In contrast most of the samples from Muleba (51.9%, n = 27/52) and Muheza(61.3%, n = 182/297) had monoclonal infections (Fws ≥0.95) (**Fig.3**). The differences in the within-host diversity of the *Pfrh5* gene infection based on the percentages of polyclonal infections among the study sites were statistically significant (p<0.001).

**Figure 3.**
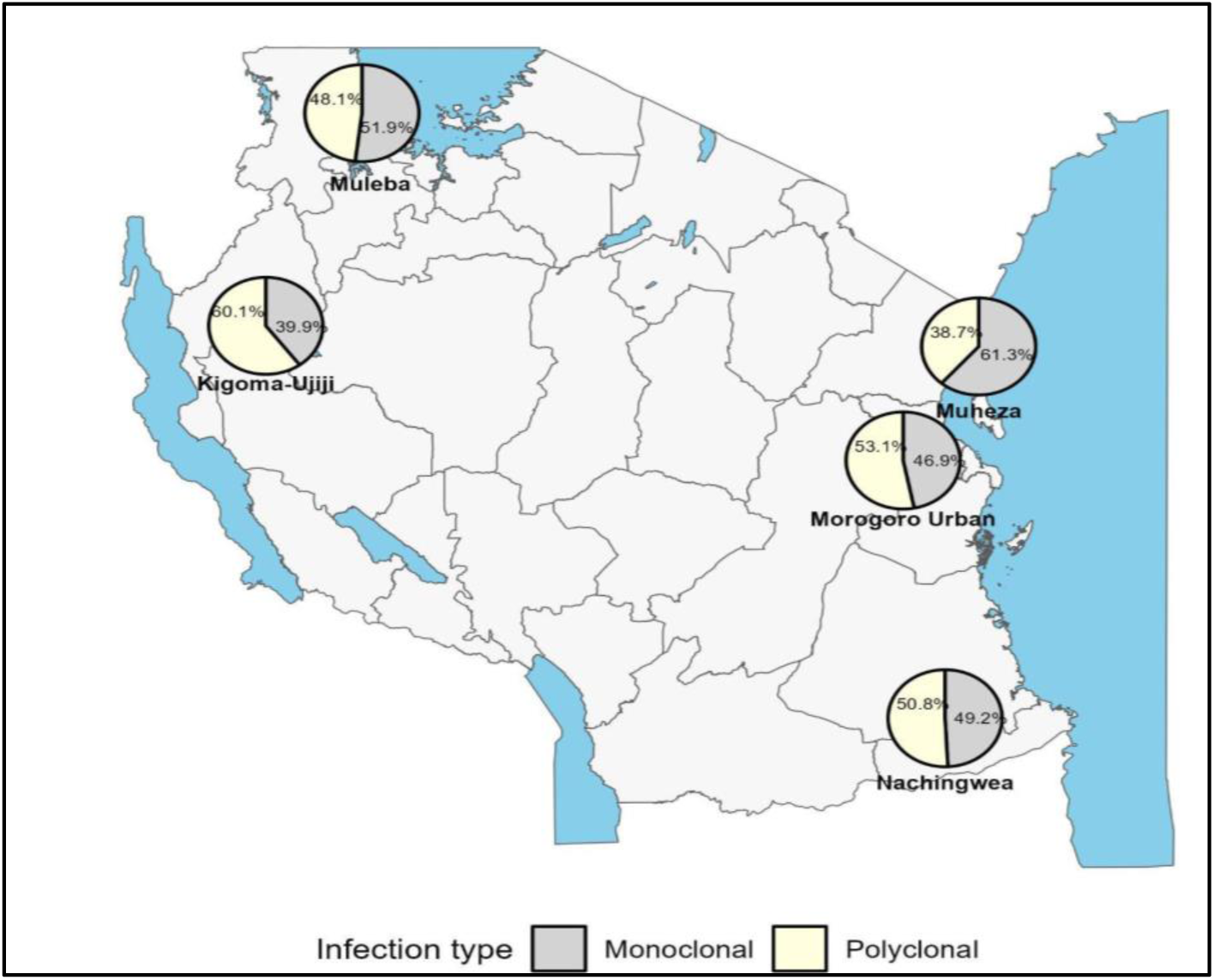
Map of Tanzania showing the percentage of monoclonal and polyclonal samples (shown by Pie charts) in study sites.

### Genetic diversity among the parasite population

The nucleotide diversity values were consistently low across all study sites ranging from 0.00053 in Muheza and Morogoro urban to 0.00068 in Nachingwea (**Fig.4**). The visualization of nucleotide diversity using violin plots showed no significant differences in levels of nucleotide diversity among the populations (**Fig.5**). Furthermore, the overall average pairwise number of nucleotide differences in the *Pfrh5* gene among sequences was 1.4, and it ranged from 1.3 in Muheza, Kigoma-Ujiji, and Morogoro urban to 1.7 in Nachingwea (**Table. 2**).

**Figure 4.**
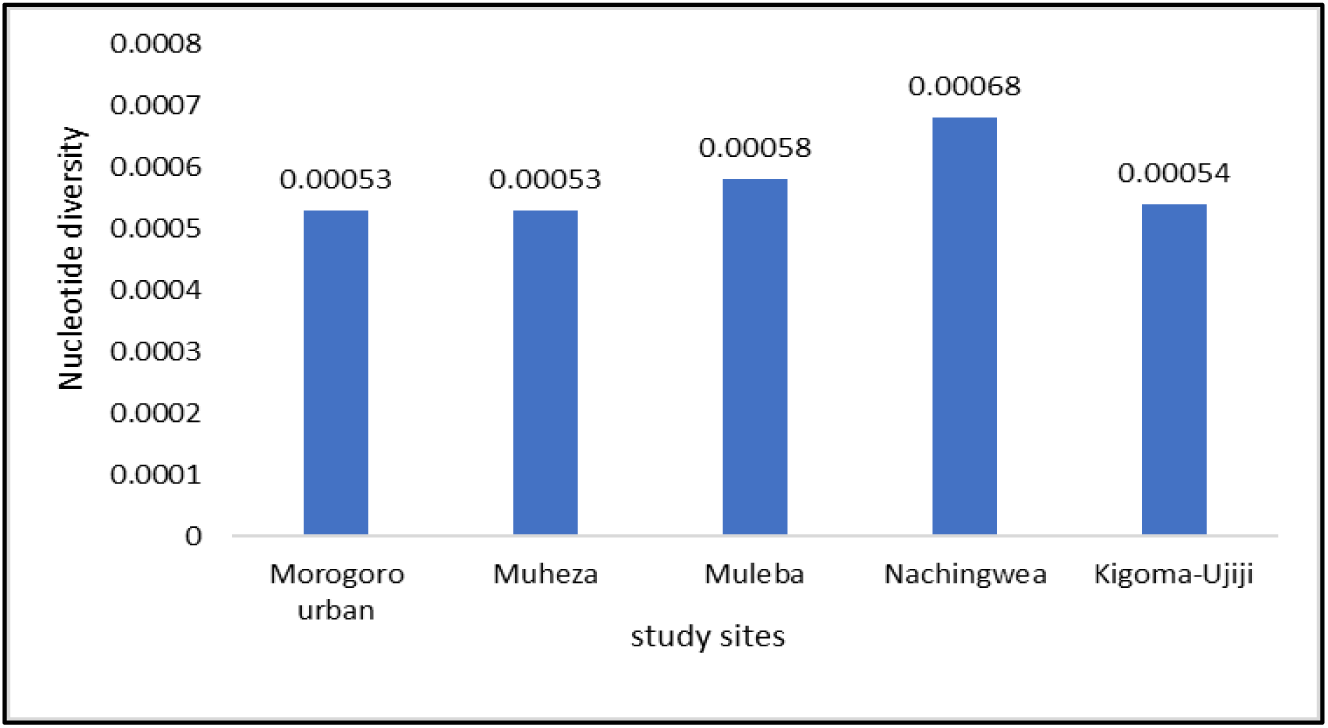
Bar plots showing nucleotide diversity in the Pfrh5 gene among the parasite populations from the five sites.

**Figure 5.**
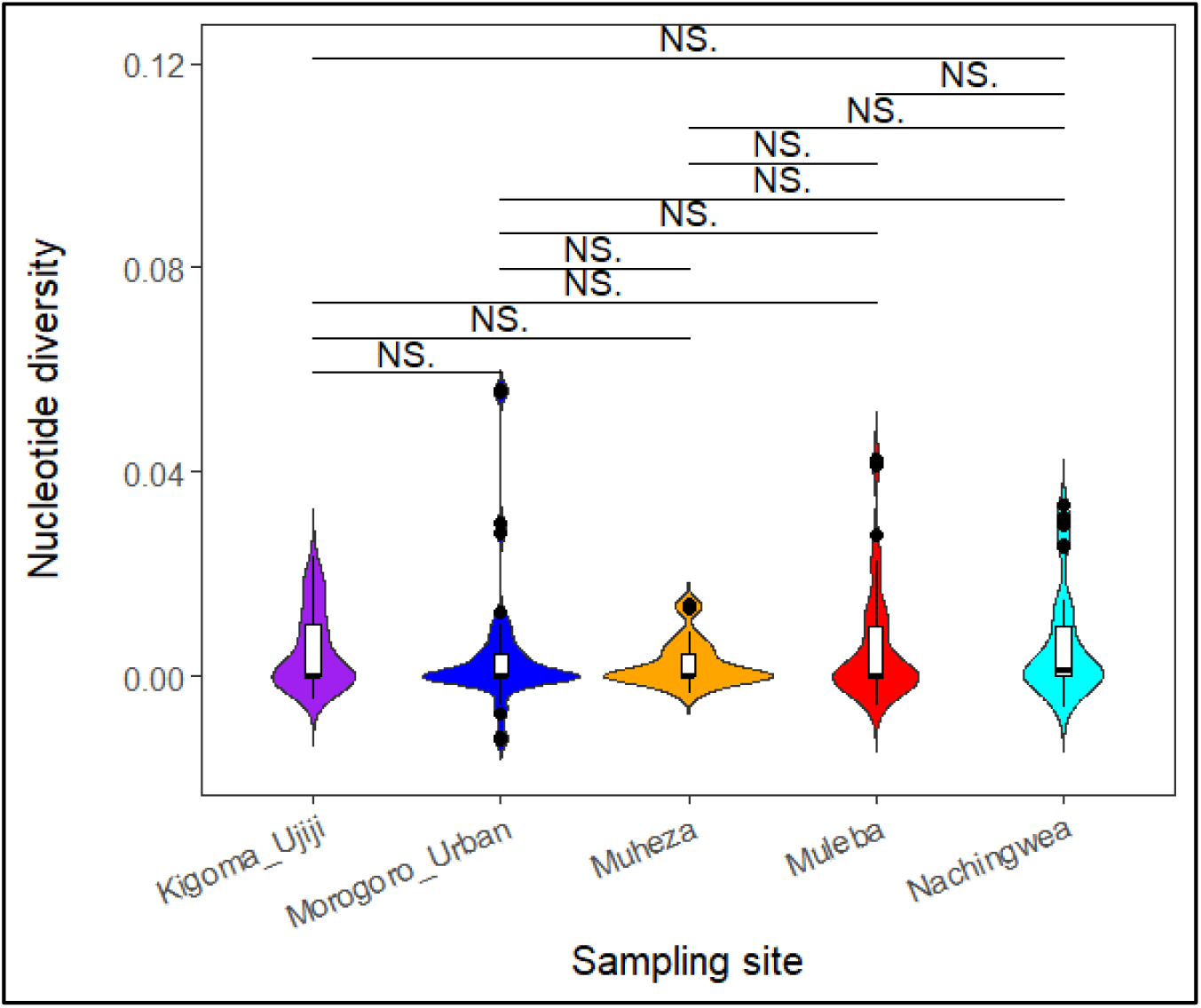
Violin plots showing the distribution of nucleotide diversity across the five populations represented with different colours. The black lines inside the box plot indicate the median nucleotide diversity. Black horizontal lines indicate the populations being compared, NS indicating no-significant difference among the populations.

**Table 2.** Measure of Pfrh5 DNA sequence polymorphisms among P.falciparum isolates in each study site.

| Polymorphisms | Study sites |  |  |  |  |
| --- | --- | --- | --- | --- | --- |
|  | Muheza | Morogoro urban | Nachingwea | Muleba | Kigoma-Ujiji |
| n | 297 | 32 | 65 | 52 | 143 |
| No. of mutations | 16 | 7 | 20 | 10 | 15 |
| No. of segregating sites | 16 | 7 | 20 | 10 | 15 |
| Synonymous mutation | 2 | 0 | 4 | 1 | 2 |
| Non synonymous mutation | 14 | 7 | 16 | 9 | 13 |
| Sn | 7 | 5 | 15 | 5 | 4 |
| Parsimony informative sites | 9 | 2 | 5 | 5 | 11 |
| k | 1.4 | 1.3 | 1.7 | 1.4 | 1.3 |
| h | 18 | 5 | 8 | 5 | 7 |
| $F_{ST}$ | 0.006 | 0.004 | 0.026 | 0.017 | 0.020 |
| Hd | 0.63 | 0.63 | 0.67 | 0.62 | 0.44 |
| Tajma's D | -1.21 | -0.74 | -1.84 | -1.02 | -1.35 |
| LD | 0.06 | 0.32 | 0.16 | 0.12 | 0.22 |
| Fu and Li's D | -2.8 | -2.3 | -4.3 | -1.7 | -0.7 |
| Fu and Li's F | -2.5 | -1.9 | -3.9 | -1.6 | -1.1 |

*n* = number of sequences, *S*- number of segregating sites, *Sn*- number of singletons, *k* = Average number of nucleotide differences, $F_{ST}$ =Wright Fixation Index, *h* = Number of haplotypes, *Hd* = Haplotype diversity, LD = Linkage disequilibrium.

To assess the extent of genetic diversity and similarity within and between the five populations, the diversity in the *Pfrh5* gene was investigated and summarized in a median-join haplotype diversity network. In total, 27 haplotypes were observed among the 313 (53.1%) monoclonal *Pfrh5* sequences. Two haplotypes, Hap_2 (n = 185) and Hap_5 (n = 71) accounted for 81.8% (n = 256/313) of the sequences, and were shared across all five regions, suggesting genetic closeness and conserved nature of the gene among these populations. Haplotype 2 was the most common *Pfrh5* haplotype, representing 59.1% (185/313) of the isolates. The sequence of Pf3D7 was identified in Hap_1, and this haplotype occurred in 16/313 (5.1%) samples, which were from Muheza(62.5%, n=10/16), Kigoma-Ujiji (18.8%, n =3/16), and Muleba (18.8%, n = 3/16) (**Fig.6)**. The haplotype diversity values (Hd) estimated among the monoclonal sequences were 0.63 in Muheza, 0.62 in Muleba, 0.63 in Morogoro urban, 0.67 in Nachingwea and 0.44, in Kigoma-Ujiji.

**Figure 6:**
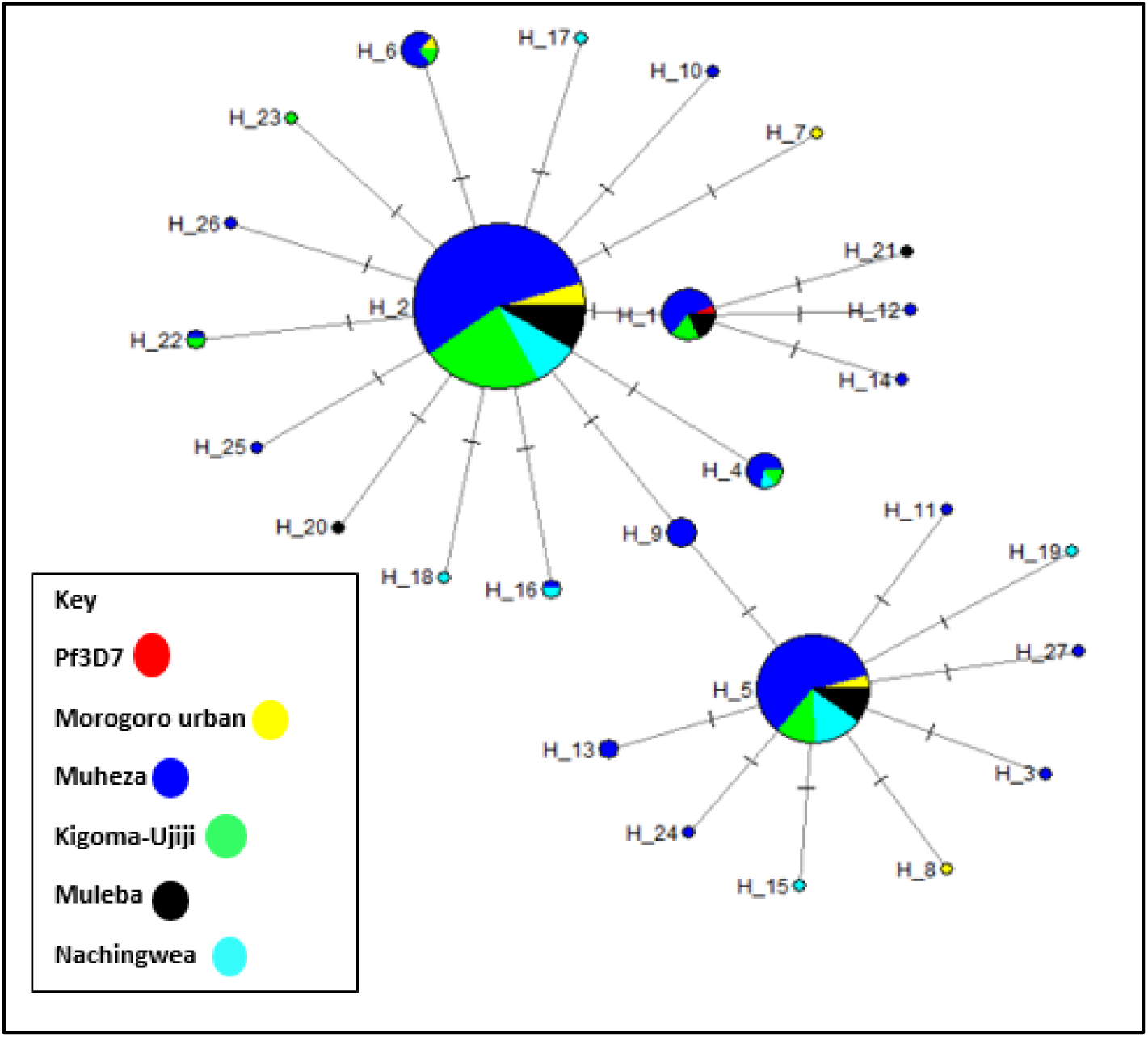
Median-Joining (MJ) haplotype network of the 27 haplotypes from Pfrh5 sequences of P. falciparum in five Tanzanian regions. The size of the circle corresponds to the number of individual samples for each haplotype, while the colours represent the geographic distribution of each haplotype.

### Population structure, differentiation and evidence of selection

Genetic differentiation estimated through the Wright Fixation Index (F_ST_) indicated that the mean F_ST_ among all sampled populations was 0.015. The F_ST_ values obtained per each site were 0.006, 0.017, 0.004, 0.026, and 0.020 in Muheza, Muleba, Morogoro urban, Nachingwea and Kigoma-Ujiji, respectively (**Table 2**). These values indicate that there is low genetic differentiation among the populations. The violin plots showed that there were no significant differences in the genetic differentiation of the *Pfrh5* gene, as estimated by F_ST_ across all the five populations (**Fig.7**). Principal component analysis did not show any population structure among parasite isolates in the sampled populations (**Fig.8a and 8b**).

**Figure 7.**
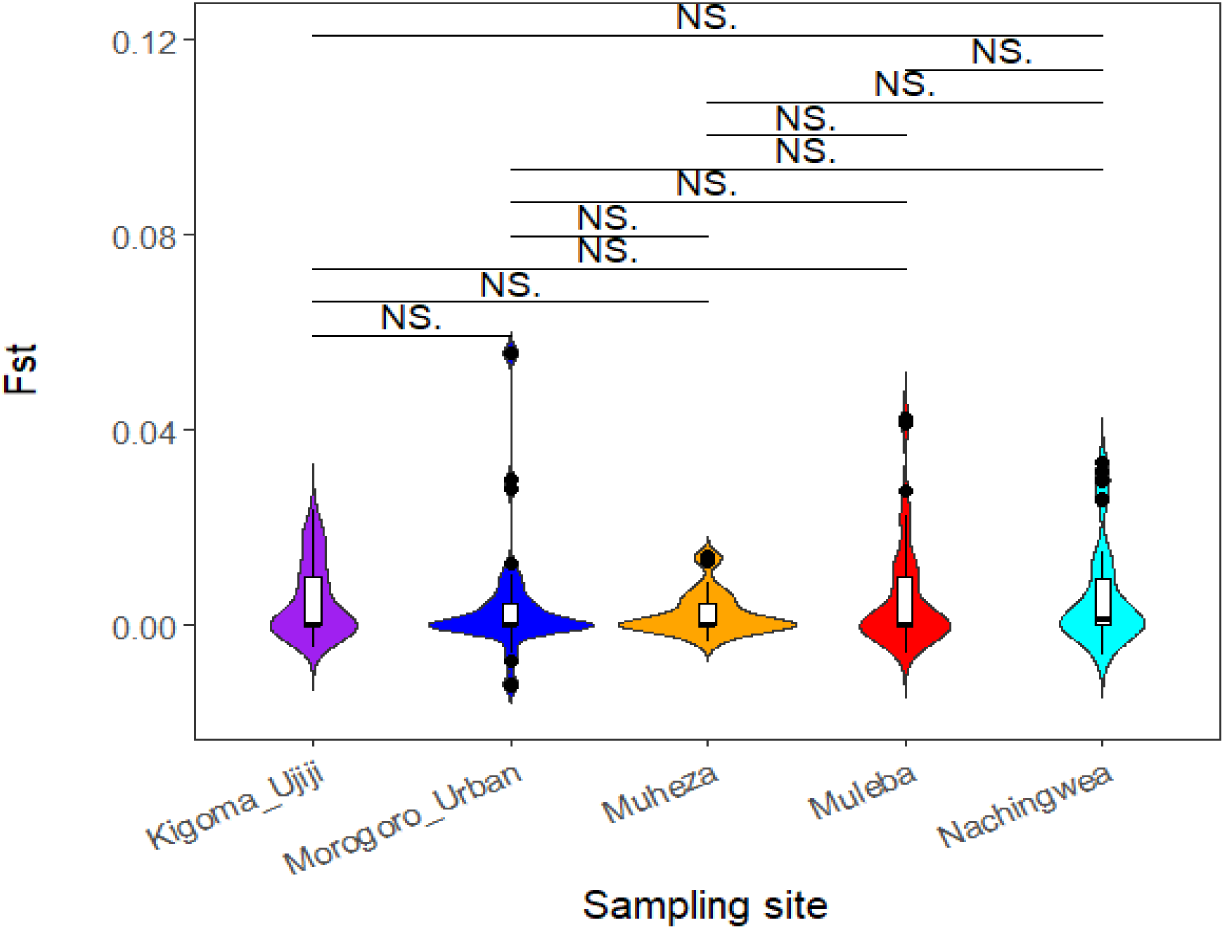
F_ST_ violin plots showing no significant difference F_ST_ values across the study populations. Black vertical lines indicate the populations which were compared, The black lines inside the box plot indicate median F_ST_ values, NS indicate no significant difference among the populations.

**Figure 8.**
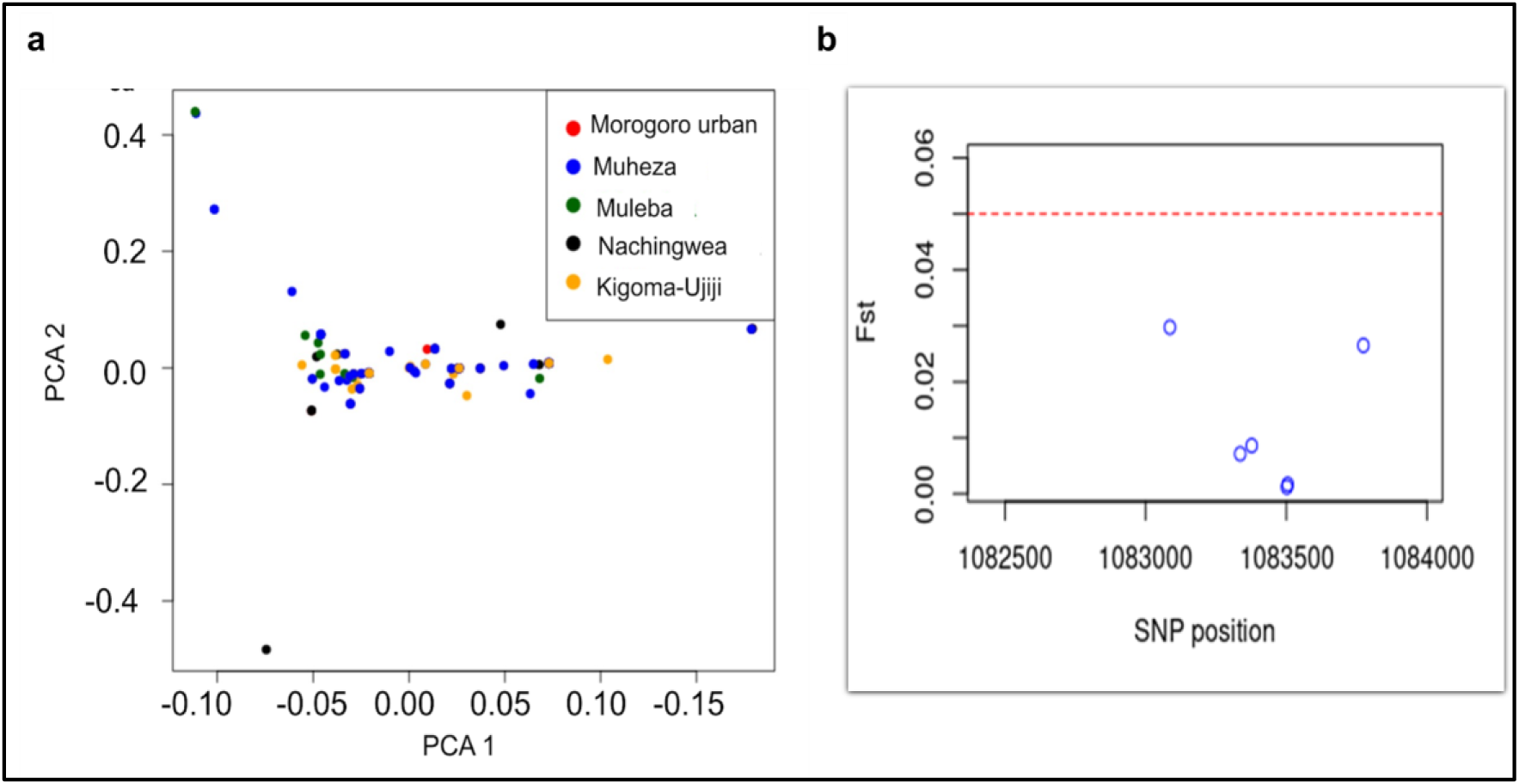
**a**. Plots of first (PC1) and second (PC2) Principal components of samples in the five districts showing no population structure. **b**)Wright’s fixation index (F_ST_) shows a borderline (red dots) value of 0.05, which indicates low genetic differentiation.

The Tajmàs D values were negative across all sites with values of −1.21 in Muheza, −0.74 in Morogoro urban, −1.84 in Nachingwea, −1.02 in Muheza and −1.35 in Kigoma-Ujiji; consistent with purifying selection patterns. The observed signatures of selection were further confirmed by the Fu and Li’s D and Fu and Li’s F test statistics where the values were negative in all the study sites (**Table 2**). The overall Linkage disequilibrium (LD) values for the *Pfrh5* gene assessing the non-random association of alleles in all the sequences was 0.11. The high LD value (0.32) was in Morogoro urban while the lowest LD value was observed in Muheza (LD = 0.06) (**Table 2**). The overall statistical values of the genetic association between polymorphic sites (Zns) and the effect of intragenic recombination on sequence polymorphism ZZ_ were 0.01 and 0.10, respectively.

## Discussion

Mainland Tanzania is classified as one of the malaria-endemic countries, with the entire population at risk of malaria infections, but with heterogeneous transmission [60]. The current pattern of malaria in Tanzania and the recent challenges attributed to epidemiological transition over the past two decades require innovative strategies to control and eventually eliminate malaria. Overall, to achieve the global malaria elimination and eradication goals, it is crucial to consider the development of an effective vaccine alongside the existing interventions. However, a significant challenge in vaccine design is the presence of polymorphisms in selected parasite proteins which render these vaccine targets ineffective even before testing them in natural populations. The *Pfrh5,* a crucial component for *P. falciparum*’s invasion of human red blood cells, has been identified as an attractive vaccine candidate since it appears essential for parasite survival [61,62]. Previous studies of the *Pfrh5* gene showed that it has limited sequence polymorphisms, and when it was used to immunize animals, it induced broadly growth-inhibitory antibodies [24,63,64]. Due to the limited data on the polymorphism of the *Pfrh5* vaccine candidate gene in Tanzania, both for standalone use and in combination with other malaria vaccine candidates, it is essential to conduct population-specific studies on the sequence diversity of *Pfrh5* to guide further development of an effective vaccine. This study evaluated the genetic diversity, population structure and identified signatures of selection within the *Pfrh5* gene from malaria parasite populations obtained from areas exhibiting varying levels of malaria transmission in Mainland Tanzania.

In this study, a measure of within infection genetic diversity (Fws) showed that the majority of samples from Muheza and Muleba had monoclonal infections as they had a high proportion of samples with Fws ≥0.95. This monoclonality reflects the high potential of inbreeding within the study populations. Higher within-host malaria parasite diversity was observed in Morogoro urban, Kigoma-Ujiji and Nachingwea where most samples had Fws < 0.95. High levels of polyclonality are indicative of a high potential for recombination and outcrossing events in the population. Additionally, high genetic diversity is typically found in high-transmission areas, where infected individuals usually carry more polyclonal infections in contrast to low-transmission areas where infections are often monoclonal [65]. The high level of polyclonality among parasites in Morogoro, Kigoma-Ujiji and Nachingwea may be due to frequent infections with multiple clones, consistent with high malaria transmission intensity. Previous studies have reported a strong positive correlation between polyclonal infections and transmission intensity [66–68].

The study observed very low nucleotide diversity across all study sites. This limited diversity likely results from the important function of the *Pfrh5* gene in the parasite’s binding and invasion of erythrocytes. Given that *Pfrh5* is essential for invasion, significant structural changes due to mutations could impair the parasite’s ability to infect red blood cells [34]. Consequently, the parasite avoids or restricts mutations in this gene, maintaining its ability to invade erythrocytes and leading to low genetic diversity. Studies also suggest that the very low values of nucleotide diversity are usually indicative of a very recent common ancestor to all sequences, as this might be expected from a hard sweep or a recent, strong bottleneck. A study of evolutionary events on human malaria suggested that a short region on chromosome 4, which encodes two essential invasion genes including the *Pfrh5* was horizontally transferred into a recent *P. falciparum* ancestor, an event that is similar to a very recent bottleneck [69]. The mean nucleotide diversity of this gene was extremely low (0.00056) compared to the nucleotide diversity seen in a well-known vaccine candidate gene, *Pfcsp* (0.0027) conducted in the same geographical areas [19]. Similar studies conducted in Mali and India also reported low nucleotide diversity in the *Pfrh5* gene [70,71].

This study also observed genetic similarity among the populations from the five sites, and a lack of population structure due to low F_ST_ value (F_ST_ < 0.05) and PCA. These findings suggest a high level of gene flow between the study populations and it is suggestive that interbreeding occurs more freely among the populations, implying the rapid spread of any introduced allele among *P. falciparum* populations. The low F_ST_ values and a lack of population structure is unlikely to be attributed to local malaria transmission within the study populations. High local transmission in these areas should have increased the likelihood of frequent sexual recombination between parasites thus increasing gene flow resulting in a more diverse parasite population. Additionally, human movements between and among the study sites which has been reported in other studies (Pereus et al, Manuscript in Preparation) should have contributed to high gene flow among the parasite population. Movements of parasites and connectedness should enable free parasite migration leading to complex parasite structure, that share some of the identical or closely related genetic features on the background of the local genomic architecture. This complex mixing of parasites may explain the lack of population structure of the *pfrh5* gene observed in this study. However, the lack of population structure could also be related to the fact that the *Pfrh5* gene is highly conserved with limited polymorphisms within and among populations [31]. Previous studies have highlighted the influence of human population mixing on promoting gene flow among *P.falciparum* isolates which likely increases the likelihood of self-fertilization and sporadic expansion of genetically identical parasites [72,73]. Also, the observed low values from other genetic metrics including nucleotide diversity, haplotype diversity and signatures of purifying selection suggest that the gene is highly conserved and these may have contributed to the low F_ST_ values and the absence of population structure among the studied populations. Similar findings of low genetic differentiation of the *Pfrh5* gene due to population structure were also reported in previous studies [74,75].

The haplotype network analysis reported 27 different haplotypes with only one haplotype identical to the Pf3D7 reference strain. Out of the 27 haplotypes, two major haplotypes were distributed across all five districts while other haplotypes were shared among sites with possible indication of the circulation of alleles across different geographical settings. Although some other haplotypes occurred independently in each site, they did not contribute significantly to the genetic diversity within the gene as the gene remains conserved as indicated by low F_ST_ values and PCA results which showed limited differentiation among isolates from the sampled population. The observed similarity of the haplotypes highlights the conserved nature of the *Pfrh5* gene, underscoring its crucial role in the parasite’s survival, particularly in erythrocyte invasion. Consequently, the limited diversity in *Pfrh5* haplotypes is beneficial for developing *Pfrh5*-based vaccines, as it suggests that a single vaccine formulation could potentially offer broad protection against diverse strains of *P. falciparum* [76]. The observed relatedness of *Pfrh5* haplotypes indicates limited genetic variability of the gene across different parasite populations. Similar findings were also observed in a study conducted in Nigeria which compared the genetic diversity of the two vaccine candidate antigens (*pfrh5* and *Plasmodium falciparum* cell traversal ookinetes and sporozoites (*Pfceltos*)). The study reported that there were low variations of haplotypes in the *Pfrh5* gene as indicated by a shorter haplotype network and low haplotype diversity compared to the *Pfceltos* gene and suggested that the *Pfrh5* gene has the potential of being a more effective subunit vaccine because of its conserved nature [77].

Tajmàs D analysis revealed negative values in all the study sites suggesting a purifying selection on this gene. This was further confirmed by the Fu and Li’s D*, and F* test statistics, where negative values were observed in all study sites using both tests. These negative population genetics summary statistics are indicative of an excess of rare variants in the *Pfrh5* gene suggesting that it is likely undergoing purifying selection and/or population expansion, both of which limits its capacity to accumulate and retain mutations. Stemming on the pivotal role of the *Pfrh5* gene on parasitès ability to invade the host cells through erythrocyte invasion, any mutations that significantly alter the structure and function of the *Pfrh5* gene could affect the parasitès ability to survive and reproduce thus these mutations are not favored, instead they are quickly eliminated in the population hence the domination of rare variants which are consistent with the observed negative Tajmàs D values. The observed results align with previous studies that reported rare variants in the *Pfrh5* gene and showed most sequences associated with a negative Tajima’s D value, suggesting a historical expansion of the parasite population [31,74]. This observation is in contrast to other malaria vaccine candidates such as the MSP1, thrombospondin-related adhesion protein (TRAP) and CSP which have been reported to exhibit a balancing selection [19,78].

Further, the assessment of non-random association of alleles at multiple sites through linkage disequilibrium revealed moderate levels of non-random association between alleles (LD = 0.1138). The limited LD is likely due to the genetic architecture of the *Pfrh5* gene which is dominated by the presence of rare variants that occur at high frequency possibly facilitated by the action of purifying selection [71]. Similar findings reporting an excess of rare variants in the *pfrh5* gene were observed in Kenya [74]. The essence of purifying selection together with the lack of population structure observed, suggests that the *Pfrh5* gene has the potential of being an effective vaccine candidate. [79]

## Conclusion

Our study assessed the genetic diversity of the *Pfrh5* gene in areas with varying levels of malaria endemicity and found the gene to be conserved, as it exhibited low nucleotide and haplotype diversity, a lack of population structure and negative Tajmàs D values as evidence of purifying selection. These findings suggest that there is high gene flow and genetic exchange within the *Pfrh5* gene. Also, the *Pfrh5* gene is under selective pressure due to its crucial role in parasite survival and may exhibit limited genetic variation across populations. This study provides important evidence on the diversity of the *Pfrh5* gene and support for the gene to be considered in the design of next generation malaria vaccines. In the future, more studies will be needed to establish if *Pfrh5* could be included in a multi-antigen vaccine targeting sporozoites, merozoites, and transmission stages, or if it could be administered alongside RTS,S. Moreover, comprehensive and intensive studies across additional sites are essential to further support the prioritization of this gene for potential inclusion in a broadly cross-protective malaria vaccine.

## Data Availability

The datasets used in this study are accessible through the MalariaGEN Plasmodium falciparum Community (Pf7k) Project.

https://www.malariagen.net/resource/34

## Availability of data and materials

The datasets used in this study are accessible through the MalariaGEN *Plasmodium falciparum* Community (Pf7k) Project https://www.malariagen.net/resource/34

## Acknowledgements

Authors thank the researchers who collected the samples and generated the WGS data, and participants and communities from the different sites where the samples and data used for this study were collected. In addition, the authors would like to thank the National Malaria Control Programme, The President’s Office, Regional Administration and Local Government, and the Regional and District authorities from the regions where the samples and data were collected and all the study participants. Additionally, the authors appreciate the support from the Bill and Melinda Gates Foundation team. Permission to publish this paper was sought and received from the Director General of the National Institute for Medical Research (NIMR).

## Ethical approval

The study utilized sequence data downloaded from the open-source MalariaGEN database (https://www.ncbi.nlm.nih.gov/pmc/articles/PMC2928508/). The study protocols for the projects that collected the samples were reviewed and approved by the Tanzanian Medical Research Coordinating Committee (MRCC) of NIMR. Permission to conduct the study in Morogoro urban-Morogoro, Muheza– Tanga, Muleba – Kagera, Nachingwea -Lindi and Kigoma-Ujiji-Kigoma was sought in writing and obtained from the district and regional medical officers. Written informed consent was obtained from patients or parents/guardians in the case of children. Appropriate information (about the study and the protocol/methods) in a language that was understood by the parents/guardians of the study patients was compiled and provided before consent was obtained. Permission to publish was obtained from the Director General of NIMR.

## Authors’ contributions

DSI- formulated the original idea, supervised data analysis and worked with AJK to write the manuscript. AJK and BML performed the analysis and wrote the manuscript with support from DP, RBM, RM and CBM. SSM, GAC, DAP, DPC and CIM reviewed and edited the manuscript. DSI revised and finalized the manuscript. All authors contributed to the article and approved the final version of the paper.

## Abbreviations

WHO: World Health Organization
SSA: Sub Saharan Africa
CSP: Circumsporozoite surface protein
MSP 1: Merozoite surface protein 1
TES: Therapeutic efficacy studies
CSS: Cross sectional studies
DDH: Designated district hospital
WGS: Whole genome sequence
VCF: Variant call format
LD: Linkage disequilibrium
PCA: Principal component analysis
PfRH5: Plasmodium falciparum reticulocyte binding homologue 5
NIMR: National Institute for Medica Research

